# EVALUATION OF THE SAFETY AND TOLERABILITY OF EXOGENOUS KETOSIS INDUCED BY ORALLY ADMINISTERED FREE BETA-HYDROXYBUTYRATE IN HEALTHY ADULT SUBJECTS

**DOI:** 10.1101/2023.06.12.23291298

**Authors:** Lisa Isabel Pimentel-Suarez, Luis Adrian Soto-Mota

## Abstract

Beta-hydroxybutyrate (D-BHB) is a metabolite with intrinsic signalling activity that has gained attention as a potentially clinically useful supplement. There are available supplements for inducing ketosis: ketone salts, ketone esters, and medium-chain triglycerides. Even when all of them raise the beta-hydroxybutyrate in the blood and all are safe and well tolerated, they significantly differ in their safety profile, their palatability, and their price. A fourth and potentially interesting option is to use biologically identical beta-hydroxybutyrate, while it is already commercially available in the United States (American Ketone LLC) and Greater China (MedPHA Ltd). However, its safety and tolerability had not yet been documented in the scientific literature. We evaluated the safety and tolerability of orally administered Free D-BHB in a gender and age-balanced sample of 24 asymptomatic and overtly healthy adults. No participant showed acid-base abnormalities or electrolyte abnormalities. Secondary symptoms were reported after only 6.2% of all drink takes and none of the reports described the symptom as “severe”. The most frequently reported secondary effects (19/720 or 2.6%) were gastrointestinal discomfort, headache (7/720 or 1%), and loss of appetite (7/720 or 1%). No correlation between weight-adjusted dose and frequency of secondary symptoms was observed. Free D-BHB was a safe and well-tolerated intervention for inducing sustained exogenous ketosis. Being bio-identical, salt-free, and lacking intermediate metabolites, this form of supplementation could have a larger safety spectrum than salt or alcohol-based exogenous ketones. More research is warranted to assess its clinical efficacy in those clinical scenarios in which achieving ketosis rapidly could be beneficial.

**KEY MESSAGES:** *What is already known on this topic:*

- Ketogenic supplements could be useful therapeutic tools in certain time-sensitive circumstances. A previously unexplored but attractive option is using bio-identical D-BHB.

*What this study adds:*

- Diluted Free D-BHB is a safe and well-tolerated intervention for inducing sustained exogenous ketosis.

*How this study might affect research, practice, or policy:*

- Being bio-identical, salt-free, further research is warranted on the potential clinical uses of Free-BHB.

## INTRODUCTION

Several ketogenic interventions have shown benefits in a wide variety of clinical scenarios [1–5]. Ketosis can be induced in different ways: by 72 hours of fasting [6], with a carbohydrate-restricted [7] diet, and within minutes using supplements [8]. While endogenous ketosis interventions have proven useful in many illnesses such as diabetes [2], psychiatric illnesses [9], migraine, [10], and cognitive decline [11], in some time-sensitive clinical scenarios where ketosis could be beneficial (for example, heart failure [12] or sepsis [13]) endogenous ketosis is impractical as patients cannot wait 72 hours for achieving a physiologically relevant production.

Beta-hydroxybutyrate (D-BHB) is a metabolite with intrinsic signalling activity [14], therefore, it has gained attention as a potentially clinically useful supplement. There are three thoroughly studied ketone supplements for inducing ketosis: ketone salts [15–17], ketone esters [18], and medium-chain triglycerides [19]. Even when they all raise the beta-hydroxybutyrate in the blood and all are safe and well tolerated, they significantly differ in their safety profile, palatability, and price [8,20].

A fourth and potentially interesting option is to use biologically identical (D-Beta-hydroxybutyrate). Being salt-free and having no intermediate metabolites or precursors, it could have a broader clinical reach than previous ketone supplements (e.g., patients with mineral load restrictions such as those living with heart failure [12] or chronic kidney disease [21,22]). While it is already commercially available in the United States (American Ketone LLC) and Greater China (MedPHA Ltd), its safety and tolerability had not yet been documented in the scientific literature. In this work, we evaluated the safety and tolerability of Free D-BHB as a ketogenic supplement in healthy adults.

## MATERIALS AND METHODS

This study was pre-registered in ClinicalTrials.gov on October 22nd, 2022, with the identifier NCT05584371. Ethical approval was obtained from the Ethics Committee of the National Institute of Medical Sciences and Nutrition Salvador Zubirán with the file code UIE-4275-22-23-1 on September 22nd, 2022. The study workflow can be observed in Figure 1.

**Figure 1.**
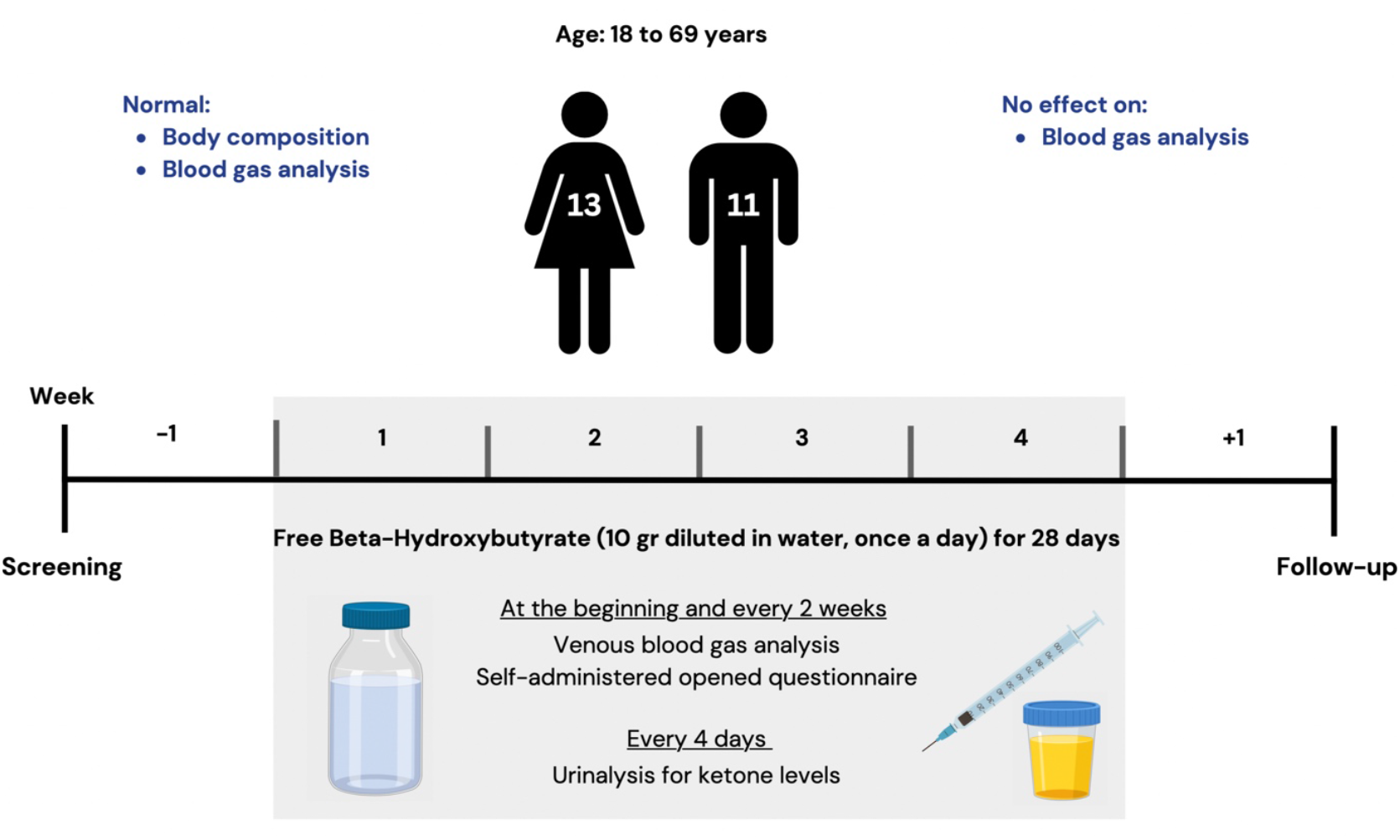
Study Workflow.

A gender and age-balanced sample of asymptomatic adults (most of them, healthcare workers) without illnesses that caused acid-base imbalances were recruited and asked to ingest 10 grams of beta-hydroxybutyrate, diluted in one litre of water and sweetened with a sugar substitute derived from the stevia plant already embedded in the preparation, every morning between 9:00 and 11:00 for four weeks. Women of childbearing age were recruited only if using a long-term method of contraception including any form of hormonal contraception and intrauterine devices. The exclusion criteria included adults on a ketogenic diet, carbohydrate-restricted diet, or intermittent fasting.

The exogenous ketones used in our study were provided by a joint donation from American Ketone LLC and MedPHA Ltd. This bio-identical D-BHB is produced with the acid hydrolysis of Poly-3Hydroxy-Butyrate [23] and is available in 60 ml bottles with 10g of free D-BHB each. Participants were instructed to dilute their content in 1 litre of water before consuming it.

As previously used in other studies on ketone supplement studies [18], the frequency and severity of symptoms (mild, moderate, severe) were assessed every two weeks with a self-administered open questionnaire to avoid priming positive and negative symptom reports and the intervention adherence was corroborated with qualitative urinary ketone strips (calculated as the proportion of urinary ketone strips with detectable acetoacetate). Venous blood gases were analysed at the baseline, 2 and 4 weeks later to rule out acid-base disorders. All blood samples were handled and analysed at the central lab of the National Institute of Medical Sciences and Nutrition Salvador Zubiran adhering to their internationally certified standard operating procedures.

Being hemodynamically stable, and consistent with previous studies, venous blood samples were preferred over arterial ones. Since acid-base analysis assumes arterial values, all acid-base variables were therefore “arterialized” by adding 0.04 to the measured pH and subtracting 5 from the measured pCO2 [24]. Because of Mexico City’s altitude, the normal range for CO2 is different than for most places in the world (30.2 ± 3.4 mmHg) [25].

Data were analysed using R version 4.0.3 with R-Studio 2022.07.2+576. Blood gas analysis was performed using CreateTableOne::CreateContTable using default parameters. Individual trajectory plots were produced using Microsoft Office Excel. The sample size was calculated for achieving a balanced age and sex sample based on previously reported safety and tolerability studies of ketone supplements [18]. All results are reported as mean +/-standard deviation and the statistical significance cut-off was p<0.05.

## RESULTS

In total 24 adults were recruited and all of them finished their participation. Their demographic characteristics by sex can be observed in Table 1.

**Table 1.** Baseline characteristics of the participants.

|  | **Women (n = 13)** | **Men (n = 11)** |
| --- | --- | --- |
| Age (years) | 42.2 ± 13.7 | 41.5 ± 14.7 |
| Height (m) | 1.6 ± 0.04 | 1.77 ± 0.04 |
| Weight (kg) | 64.3 ± 8.8 | 78.9 ± 14.8 |
| Dose (mg/kg) | 158 ± 20.9 | 129.8 ± 19.2 |
| Body Mass Index | 23.9 ± 3.2 | 20.9 ± 6.3 |

\* Data are means ± standard deviation.

No participant showed abnormalities in their vital signs or acid-base assessments in any of their visits (Figure 2). Additionally, all acid-base parameters were non-significantly different across all time points (Table 2). Additionally, no electrolyte abnormalities in Na, K, Cl, or ionized Ca were observed (shown in the publicly available dataset).

**Table 2.** Acid-base analyses across all time points (arterialized values).

|  | **Visit 1** | **Visit 2** | **Visit 3** | **p-value** |
| --- | --- | --- | --- | --- |
| pH | 7.41 ± 0.03 | 7.43 ± 0.03 | 7.44 ± 0.03 | 0.19 |
| pCO<sub>2</sub> | 39.2 ± 4.3 | 38.5 ± 5.8 | 37 ± 4.7 | 0.30 |
| HCO<sub>3</sub><sup>-</sup> | 24.9 ± 2.6 | 24.8 ± 2.3 | 24.4 ± 1.9 | 0.72 |

\* Data are means ± standard deviation. Hypothesis testing used a Kruskal-Wallis Test.

**Figure 2.**
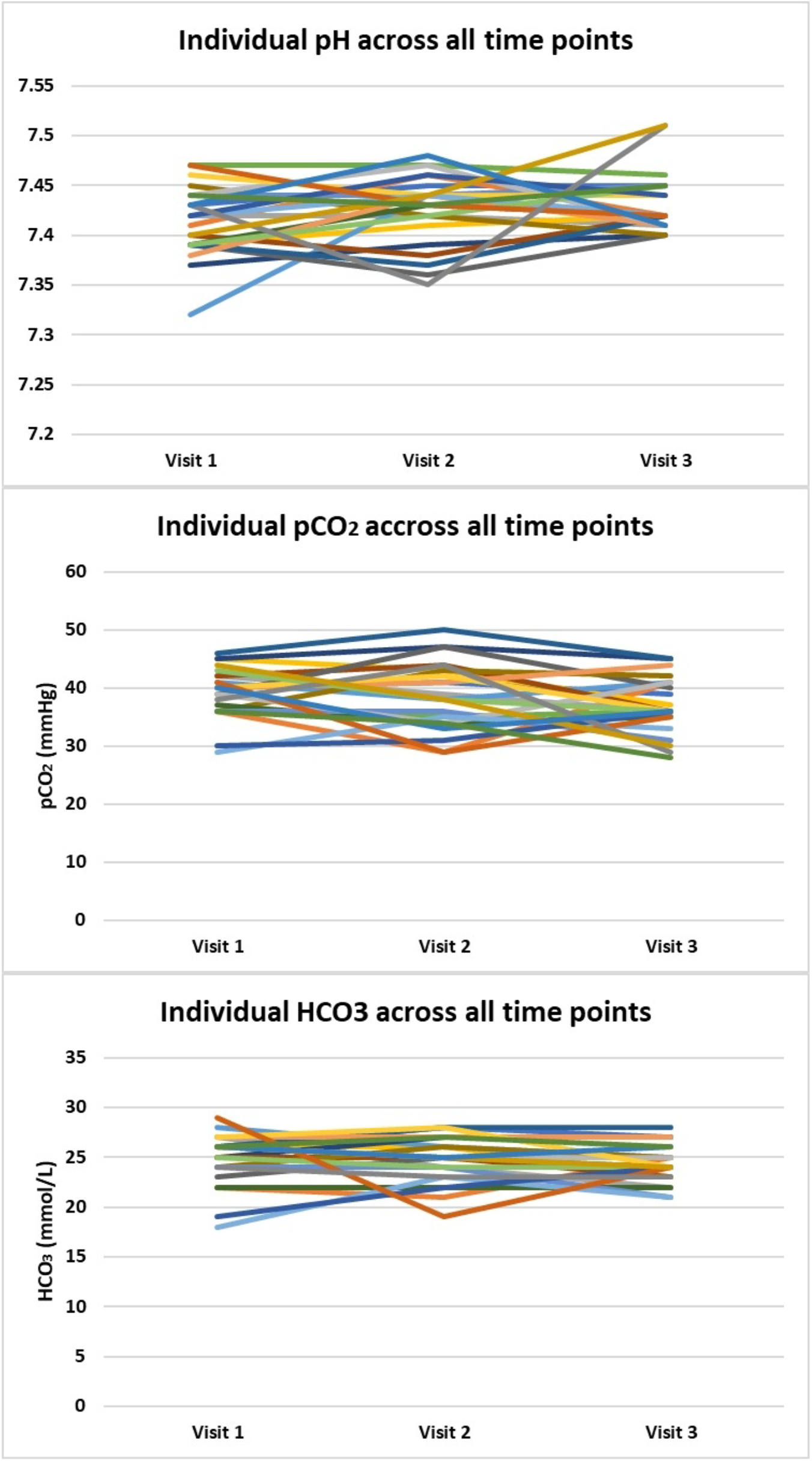
The individual trajectory of acid-base parameters across all time points. Panel A) pH. Panel B) pCO2. Panel C) HCO-3.

Regarding tolerability and palatability, all participants responded to the self-reported questionnaires at least once and had an adherence above 90%. Only 44 out of the 720 total takes (6.2%) of the drink reported secondary symptoms. Of which, none of the reports described the symptom as “severe”, (17/720 or 2.3%) were “moderate” and the remaining (27/720 or 3.9%) were “mild” (Table 3).

**Table 3.** Secondary symptoms following each of the 720 administered ketone drinks.

| **Symptom** | **No of participants** | **Events\***<br>**(mild/moderate)** |
| --- | --- | --- |
| Gastrointestinal discomfort | 8 | 20/720 (10/10) |
| Loss of appetite | 5 | 12/720 (9/3) |
| Headache | 6 | 12/720 (8/4) |

\* Some participants recorded the same symptom more than once, so the occurrence was expressed relative to 720, the total number of drinks consumed (24 x 30 drinks).

The most reported secondary effects (19/720 or 2.6%) were gastrointestinal discomfort, headache (7/720 or 1%), and loss of appetite (7/720 or 1%). The rest of the reported secondary symptoms were reported <1% of the time. No participant reported more than 2 secondary symptoms nor the same symptom more than twice. No correlation between weight-adjusted dose and frequency of secondary symptoms was observed (Figure 3).

**Figure 3.**
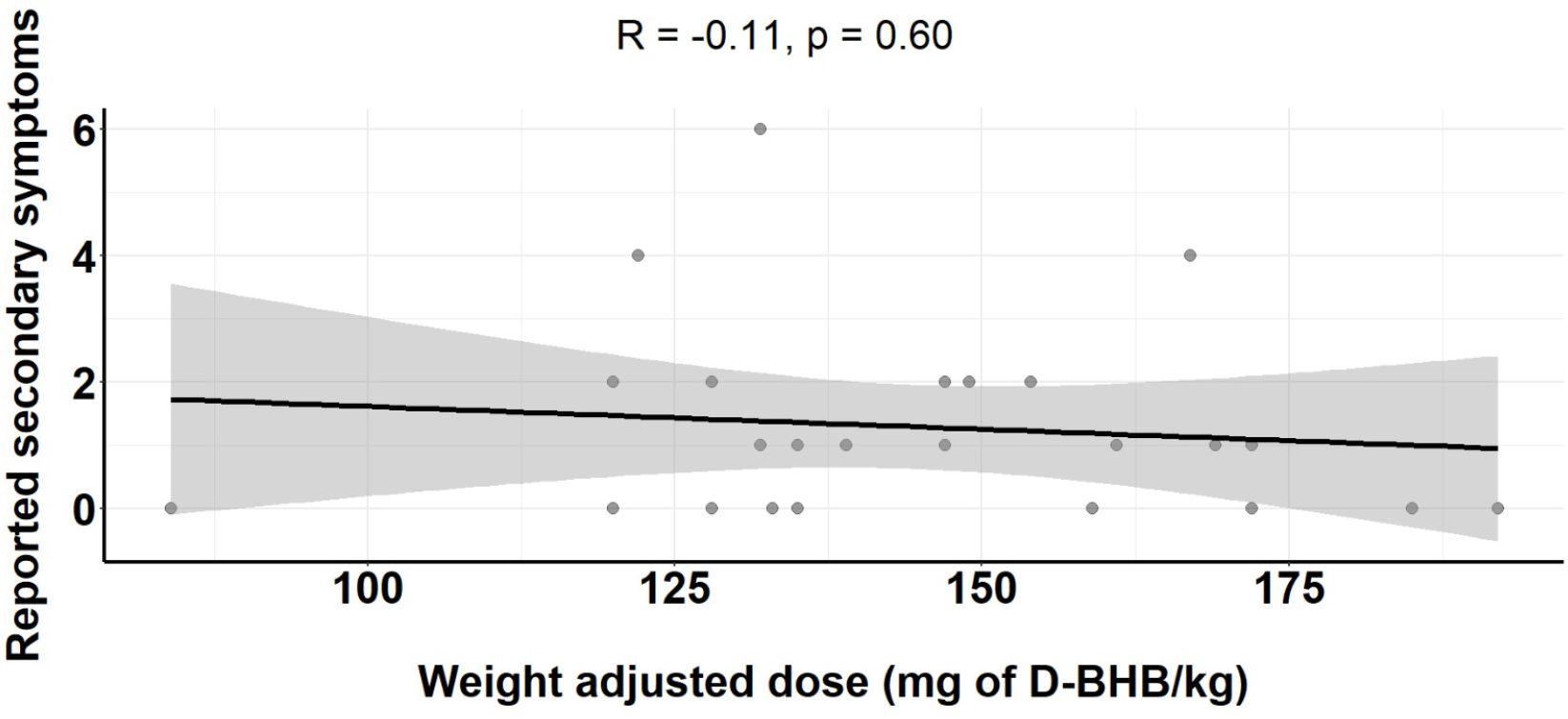
Correlation between secondary symptom frequency and weight-adjusted dose.

## DISCUSSION

Hereby and as observed with other formulations [16,18], exogenous ketosis with Free D-BHB was safe and well tolerated. Since Free-D-BHB has some advantages over other formulations, more research on its potential clinical applications is warranted.

For example, ketone salts may not be suitable for patients who should restrict their salt intake, such as those with heart failure or kidney disease [26]. On the other hand, the ketone monoester has an alcohol precursor that could limit its use in patients with liver disease and has poor palatability [3,20]. More importantly, being bio-identical, it is reasonable to assume that the safety and pharmacokinetic properties of Free D-BHB are comparable to the endogenously produced metabolite.

It is worth reminding that since Mexico City is more than 2200 meters above sea level, the geometrical normal values are slightly different, and it is expected to observe lower CO2 and HCO3 levels in healthy individuals. This difference is relevant because clinicians elsewhere could incorrectly diagnose respiratory alkalosis with CO2 values that are normal for people living in Mexico City. [25] Consistently with other ketone supplement formulations, even those inducing acute documented acid-base disturbances [8], the chronic ingestion of Free D-BHB did not cause chronic acid-base or electrolyte alterations [8,15,16,18].

Among the strengths of this study, we should mention that most participants were healthcare workers and adherence was above 90%, making it unlikely that the low frequency of adverse effects is due to lack of adherence, and the close communication with participants. Moreover, since all participants consumed the same dose but have different weights a dose/kg spectrum is created allowing us to see if the frequency of adverse symptoms is more frequent in leaner individuals. As with other ketogenic supplements, no correlation was observed [18].

Furthermore, we examined a sample size that is comparable to what has been examined for other commercially available ketogenic compounds. However, in contrast with those studies, we were able to enlist a more diverse age and gender distribution in our study. For example, Holland et al [15], who only studied individuals between the ages of 18 and 35, and Stefan et al [17], focused exclusively on teenagers. This difference is particularly relevant as some of the potential clinical applications of exogenous ketones like heart failure [12] or cognitive decline [11] are on illnesses that are more frequent among elders.

Acid-base and blood electrolytes are the only markers that different types of exogenous ketones have shown to induce changes on [8]. Therefore, when comparing studies evaluating the safety of different exogenous ketones, it is important to consider that some of the relevant markers, they measured are specific to the chemical nature of each supplement. In other words, due to their alcohol content, liver enzyme measurements are relevant for butanediol or the ketone ester but not for bioidentical BHB. Moreover, despite having used BHB doses many times greater than the one we worked with, none of the exogenous ketone supplements has been shown to induce detrimental changes in liver or kidney function tests [8,10,15,16,18]. Notably, the only case where other biomarkers have been shown to change after consuming a ketone supplement, are migraine patients with baseline metabolic risk markers, in whom these markers (i.e. C-reactive protein, cortisol insulin and thyroid markers) improved [10].

On the other hand, although comparable to what has been documented with other forms of ketone supplementation [8,10,15,16,18], we should mention its follow-up time among its limitations. Additionally, is not possible to make direct comparisons about palatability between the other available ketogenic supplements, and more studies directly addressing this question are required [20].

## CONCLUSIONS

Diluted Free D-BHB was a safe and well-tolerated intervention for inducing sustained exogenous ketosis. Being bio-identical, salt-free, and lacking intermediate metabolites, this form of supplementation has a larger safety spectrum than salt or alcohol-based exogenous ketones. More research is warranted to assess its clinical efficacy in those clinical scenarios where achieving ketosis rapidly could be beneficial.

## Data Availability

Anonymized data and analysis code are publicly available at: https://github.com/AdrianSotoM/FreeBHB.

## DECLARATIONS

### Funding

The exogenous ketones used in our study were provided by a joint donation from American Ketone LLC and MedPHA Ltd. However, the authors designed, carried out, analysed all data and wrote this report independently.

## Acknowledgements

The authors thank the Central Laboratory staff at the National Institute of Medical Sciences and Nutrition Salvador Zubiran for their invaluable support to make this work possible.

## Conflicts of Interest

Both authors declare they are free of any financial interests or personal relationships that could have influenced their interpretation or reporting of these data.

## CONTRIBUTORSHIP STATEMENT

LIPS recruited participants and collected data, and ASM designed the study and analysed data. Both authors redacted this draft and approved the current version of the manuscript.

## Notes

### Competing Interest Statement

The authors have declared no competing interest.

### Clinical Trial

NCT05584371

### Author Declarations

Ethics Committee of the National Institute of Medical Sciences and Nutrition Salvador Zubiran gave ethical approval for this work with the file code UIE-4275-22-23-1 on September 22nd, 2022.

